# From Data to Insights: A Tool for Comprehensive Quantification of Continuous Glucose Monitoring (QoCGM)

**DOI:** 10.1101/2025.01.01.25319870

**Authors:** Simon Lebech Cichosz, Stine Hangaard, Thomas Kronborg, Peter Vestergaard, Morten Hasselstrøm Jensen

**Author notes:** Corresponding author: Simon Cichosz, Postal address: Selma Lagerløfs Vej 249, 12-02-048, 9260 Gistrup, Danmark.

## Abstract

Continuous Glucose Monitoring (CGM) has become a important technology in the management and research of both type 1 and type 2 diabetes, providing real-time data on glucose fluctuations that were previously inaccessible with traditional monitoring methods. Numerous analytical tools, such as cgmquantify, iglu, GLU, rGV, and CGManalyzer, have been developed for platforms like R and Python to calculate standard metrics and extract insights from CGM data. However, these tools often fail to address the full spectrum of analytical requirements. Furthermore, there is a significant lack of updated, open-source tools tailored for MATLAB—a platform widely used by the research community. To address this gap, we introduce Quantification of Continuous Glucose Monitoring (QoCGM), a comprehensive, open-source analytical tool for CGM data specifically designed for MATLAB. A case study involving 324 individuals with insulin-treated type 2 diabetes mellitus (T2DM) demonstrates the utility of QoCGM, highlighting the distinct aspects of glucose dynamics captured by different CGM-derived metrics through an analysis of their coefficients of determination (R²).

## Introduction

Continuous Glucose Monitoring (CGM) has emerged as a cornerstone in the management and study of both type 1 and type 2 diabetes [1–4], offering real-time insights into glucose fluctuations that were previously unattainable through traditional monitoring methods. The data generated by CGM devices provides an unprecedented opportunity for both clinical and scientific applications, enabling precise glycemic control, better prediction of hypoglycemic and hyperglycemic events, and a deeper understanding of glucose dynamics. Key metrics derived from CGM data, such as time-in-ranges and glycemic variability have become integral to both patient care and diabetes research, facilitating personalized treatment plans and the development of novel therapeutic strategies [5,6].

In several previous studies, we have shown how CGM derived metrics can be utilized in the prediction of emerging hypoglycemic events [7–10], week-to-week risk of excessive hyperglycemia, hypoglycemia and glycemic variability [11], elevated ketone levels, [12] and identification of gastroparesis [13].

Over the years, several analytical tools have been developed to harness the potential of CGM data, providing capabilities for calculating standard metrics and generating insights from the complex glucose profiles captured [14]. Examples of such tool are cgmquantify, iglu, GLU, rGV, and CGManalyzer, which are all available for R or python [15–18]. However, these tools do often not address the full spectrum of analytical needs. Specifically, there is a growing demand for tools that can incorporate newer, clinically relevant metrics, perform nuanced analyses of nocturnal versus diurnal glucose patterns, event detection of hypoglycemia, and assess day-to-day variations in glucose levels—factors that are critical for both advanced research and personalized clinical interventions. Moreover, while various software solutions exist, there is a notable gap in the availability of updated, open-source tools for the MATLAB environment, which is widely used in the Engineering research community.

This article presents an open-source tool Quantification of Continuous Glucose Monitoring (QoCGM) designed for comprehensive CGM data analysis within the MATLAB environment. This tool does not only include newer metrics but also offers features for examining night/day glucose variations and day-to-day changes, making it a valuable resource for researchers and clinicians.

## Implementation

QoCGM is implemented in MATLAB and has been tested with version R2021b. The source code is available on GitHub (https://github.com/simcich/QoCGM). QoCGM is designed to process preprocessed CGM data from comma-separated value (CSV) files. The input CSV files should contain two columns: one for the timestamp in the format ‘YYYY-MM-DD HH:MM:SS’, and one for the corresponding glucose values, which can be in either mg/dL or mmol/L.

The processing of CGM data in QoCGM occurs in two primary stages: (i) preprocessing and handling missing data, and (ii) deriving metrics. The source code includes examples that demonstrate how to use QoCGM with a single file or to batch process multiple files within a directory.

To run QoCGM, users load the CGM data into a table and provide the following additional arguments:

- Sampling frequency: A numeric value representing the expected sampling frequency in minutes. For example, set this to 5 if measurements are expected every 5 minutes.
- Sampling tolerance: A numeric threshold (in minutes) defining the acceptable variation in sampling times. Measurements that fall outside this threshold are corrected.
- End of night: An integer (0-24) specifying the hour at which the ‘morning’ period begins, thereby marking the end of the ‘night’. For instance, set this to 6 for a 6 AM start.
- Conversion flag: A binary flag (1 or 0) indicating whether to convert CGM values from mmol/L to mg/dL. Since the metrics are derived based on mg/dL values, conversion is necessary if the CGM units are in mmol/L.
- Plotting flag: A binary flag, set to 1 to enable plotting of figures related to signal quality control and data processing.
- Handling of missing data: either interpolation (default) or by removing periods with missing data.

Upon execution, QoCGM generates a CSV file containing the derived CGM metrics, which can then be imported into statistical software for further analysis.

## Preprocessing and Handling Missing Data

### Data Preprocessing and Removal of Duplicates

The preprocessing of the CGM signal begins with the removal of duplicate entries in the dataset, specifically focusing on the timestamps. This step is critical to ensure that each time point in the data is unique, preventing potential errors in subsequent analyses.

### Identification of Missing Periods in the CGM Signal

CGM systems are configured to sample glucose levels at specific intervals. For example, a common sampling frequency involve recording data every 5 minutes. This interval is considered the expected time between consecutive measurements.

Small deviations from the expected sampling interval can occur due to natural variability in device performance or other factors. To account for this, a tolerance range is established around the expected interval, defining the minimum and maximum allowable intervals between consecutive readings. The time differences between consecutive glucose measurements are calculated to identify intervals that fall outside the permissible % range. Any interval that is shorter or longer than the defined tolerance indicates a potential gap or missing period in the data. The specific points in time where data is missing are identified by examining the calculated time differences. These points mark the beginning of periods where the expected glucose measurements are absent.

### Interpolation of Missing Data

To reconstruct the missing portions of the CGM signal, interpolation is performed over the identified gaps using Piecewise Cubic Hermite Interpolating Polynomial (PCHIP) [19].

The final PCHIP interpolating function H(x) is defined piecewise over each interval [x_i_,x_i+1_] as:

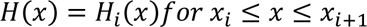

This method was chosen for its ability to maintain the shape and smoothness of the original data, making it suitable for CGM signals, which typically exhibit smooth and continuous changes in glucose levels. This reconstructed signal filled the gaps identified earlier, providing a continuous and complete signal for further analysis. *Figure 1* illustrates a CGM signal with missing periods, the raw signal, and the PCHIP interpolation. It is also possible to handle missing periods without interpolation by setting input argument to removing these periods.

**Figure 1.**
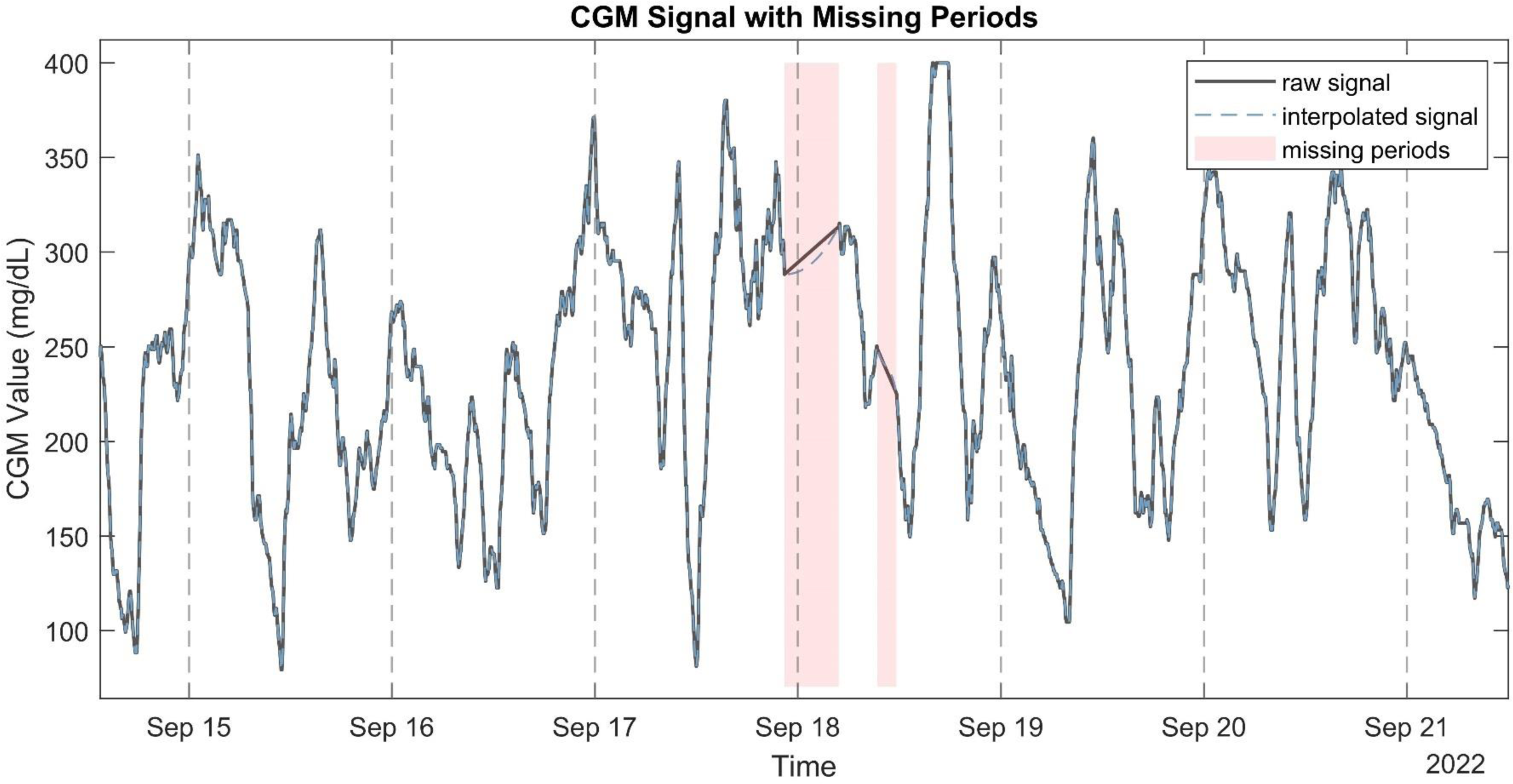
example of an raw CGM signal with the corresponding interpolated signal and missing periods highlighted.

## CGM Derived Metrics

After preprocessing and interpolation of the signal, QoCGM derives a set of CGM metrics, listed in *Table 1*. Glucose metrics output by QoCGM were chosen to represent categories of glucose characteristics that reflect a set of broad domains that relate to outcomes, glycemic control, or be influenced by exposures in clinical trials.

**Table 1.**
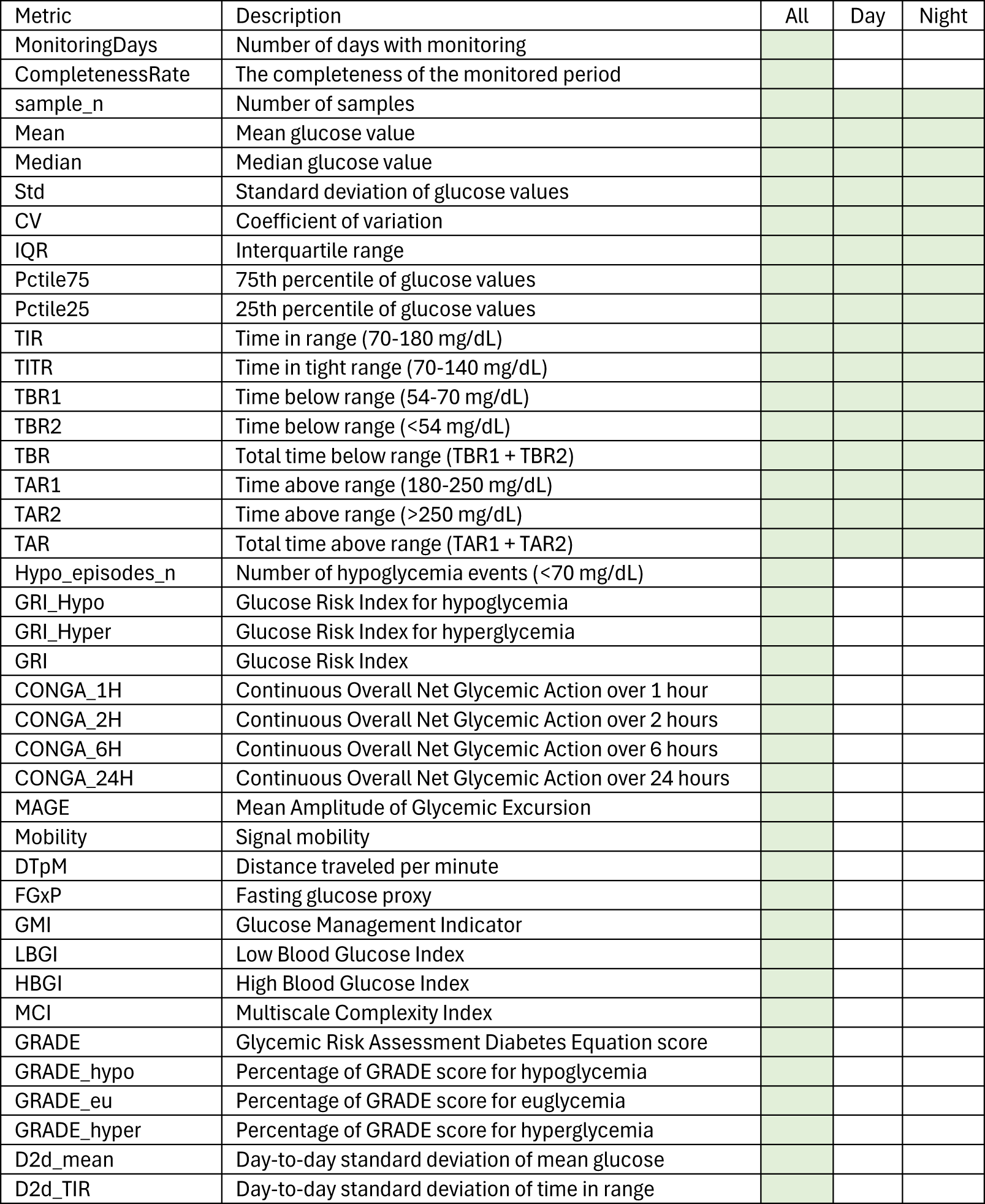
The figure displays the CGM metrics calculated by QoCGM with a short description.

The domains are:

- Basic descriptive statistics
- Time-in-Range (TIR) metrics
- Glycemic risk indicators
- Glycemic variability metrics
- Glycemic control indicators
- Entropy and complexity measures

The QoCGM tool calculates Basic Descriptive Statistics and Time-in-Range (TIR) Metrics for the entire day, as well as separately for nighttime and daytime periods. Additionally, it provides the day-to-day standard deviation for selected variables. All other metrics are derived from the complete CGM signal provided.

QoCGM aggregates the average of each summary variable across all days for each participant, resulting in a single overall value for each metric. For instance, QoCGM reports TIR metrics such as: (i) mean TIR for whole days, (ii) TIR for nighttime based on a predefined interval, (iii) TIR for daytime based on a predefined interval, and (iv) day-to-day variation in TIR. Metrics for each day for a given individual can be calculated by splitting the CGM signal into separate files corresponding to unique days.

### Basic Descriptive Statistics

We included a range of basic descriptive statistics to characterize the CGM data for each participant. These metrics provide foundational insights into the central tendency, dispersion, and distribution of glucose values over the monitored period.

- Mean Glucose: The average glucose level over the monitoring period.
- Median Glucose: The midpoint of the glucose distribution, less influenced by outliers.
- Standard Deviation (SD): A measure of glucose variability around the mean.
- Coefficient of Variation (CV): The standard deviation normalized by the mean, expressed as a percentage.
- Interquartile Range (IQR): The range between the 25th and 75th percentiles, representing the spread of the middle 50% of glucose values.
- 75th and 25th Percentiles: Indicators of the upper and lower bounds of the middle half of the glucose distribution.

These metrics were calculated for the entire period, as well as separately for daytime and nighttime, to capture diurnal variations in glucose levels.

### Time-in-Range (TIR) Metrics

TIR metrics were employed to assess the proportion of time that glucose levels remained within clinically relevant thresholds, providing a detailed view of glycemic control. The following TIR metrics were included in the analysis:

- TIR (70-180 mg/dL) [5]: Percentage of time within the target range, indicating overall glycemic control.
- TITR (70-140 mg/dL) [20]: Percentage of time within a narrower target range, reflecting tighter glucose control.
- TBR1 (54-70 mg/dL) [5]: Percentage of time in level 1 hypoglycemia.
- TBR2 (<54 mg/dL) [5]: Percentage of time in level 2 hypoglycemia.
- Total TBR [21]: Combined percentage of time in TBR1 and TBR2.
- TAR1 (180-250 mg/dL) [5]: Percentage of time in level 1 hyperglycemia.
- TAR2 (>250 mg/dL) [5]: Percentage of time in level 2 hyperglycemia.
- Total TAR [5]: Combined percentage of time in TAR1 and TAR2.

These metrics were calculated for the whole day and separately for daytime and nighttime.

### Glycemic Risk Indicators

Glycemic Risk Indicators were calculated to quantify the risk associated with hyperglycemia and hypoglycemia, providing a nuanced assessment of the overall glycemic profile. The following metrics were included from the Glycemic Risk Index (GRI), Low/High Blood Glucose Index (LBGI/HBGI), and The Glycemic Risk Assessment in Diabetes (GRADE):

- Glycemic Risk Index for Hypoglycemia (GRI Hypo) [22]: This metric quantifies the risk associated with hypoglycemic episodes, with higher values indicating a greater risk of low glucose levels.
- Glycemic Risk Index for Hyperglycemia (GRI Hyper) [22]: This metric assesses the risk of hyperglycemia, with higher values indicating a greater risk of elevated glucose levels.
- Overall Glycemic Risk Index (GRI) [22]: A composite metric combining the risks of both hypoglycemia and hyperglycemia, calculated as a weighted sum of GRI Hypo and GRI Hyper.
- Low Blood Glucose Index (LBGI) [23]: This score measures the risk of experiencing low blood glucose levels, with higher values indicating increased hypoglycemia risk.
- High Blood Glucose Index (HBGI) [24]: This score quantifies the risk of high blood glucose levels, with higher values indicating an increased risk of hyperglycemia.
- GRADE (Overall) [21]: This metric provides a summary measure of glycemic risk across all glucose ranges, combining the risks of hypoglycemia, euglycemia, and hyperglycemia.
- GRADE for Hypoglycemia [21]: The specific component of the GRADE metric that assesses the risk of hypoglycemia.
- GRADE for Euglycemia [21]: The component of the GRADE metric that evaluates the time spent in the euglycemic range (normal glucose levels).
- GRADE for Hyperglycemia [21]: The component of the GRADE metric that quantifies the risk associated with hyperglycemia.

These glycemic risk indicators were calculated based on the entire CGM signal, offering a detailed view of the participant’s glycemic risk profile. A detailed mathematical definition of the metrics is provided in *Supplementary Material S1*.

### Glycemic Variability Metrics

Glycemic Variability Metrics were utilized to assess fluctuations in glucose levels over time, providing insights into the stability and predictability of glycemic control. The following metrics were included:

- CONGA (Continuous Overall Net Glycemic Action) [25]: This metric measures glucose variability over different time intervals. We calculated CONGA for 1-hour, 2-hour, 6-hour, and 24-hour periods (CONGA 1H, CONGA 2H, CONGA 6H, and CONGA 24H), reflecting short-term to daily fluctuations in glucose levels.
- MAGE (Mean Amplitude of Glycemic Excursions) [26]: MAGE captures the average magnitude of significant glucose swings, both increases and decreases, by focusing on excursions that exceed one standard deviation from the mean. It is a widely used indicator of glycemic variability and the likelihood of large glucose fluctuations.
- Mobility: This metric assesses the signal mobility by measuring the variance of glucose changes (differences between consecutive glucose readings) relative to the overall variance of the glucose signal. Higher mobility indicates greater variability in glucose levels. [27]
- Distance Traveled per Minute (DTpM) [28]: DTpM quantifies the total amount of glucose fluctuation over time by summing the absolute changes in glucose levels and normalizing by the total duration of monitoring. This metric provides a rate of glucose change, highlighting periods of rapid fluctuations.
- Standard Deviation Day-to-Day (SD d2d): This metric measures the day-to-day variability in glucose levels, specifically assessing the mean glucose and Time-in-Range (TIR) variability across days. It captures the consistency of glycemic control from one day to the next.

These Glycemic Variability Metrics provide a comprehensive understanding of the dynamics of glucose levels, identifying periods of instability that may increase the risk of hypo- or hyperglycemic events and contribute to long-term complications in diabetes management. A detailed mathematical definition of the metrics is provided in *Supplementary Material S1*. For example, in *Figure 2*, the peaks and nadirs above σ (1 SD) is highlighted in the CGM trace. The absolute excursions |λ|, both positive and negatives, are used to calculate the MAGE metric as show in the equation below:

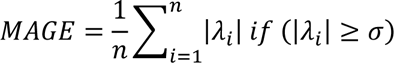

**Figure 2.**
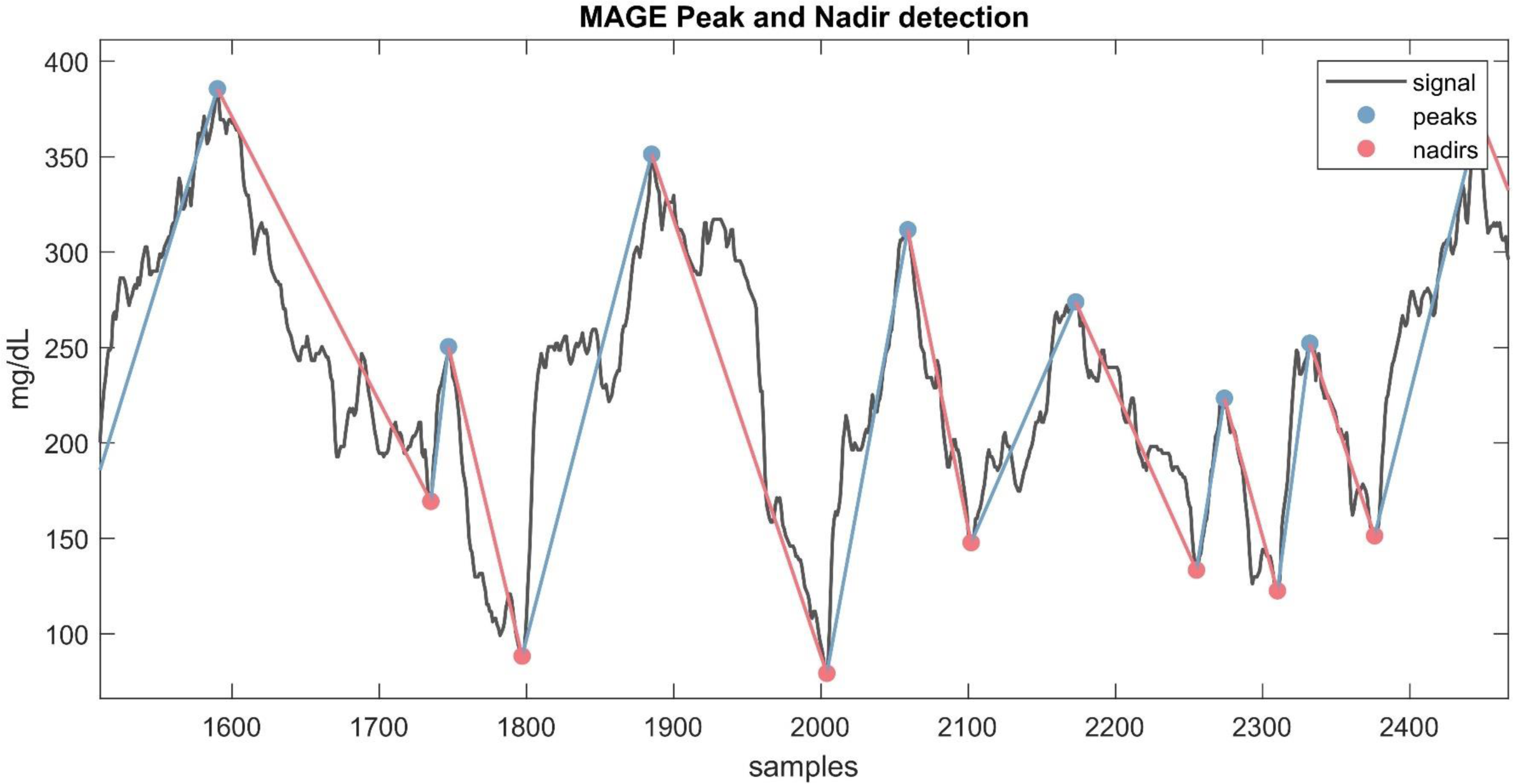
example of an CGM signal with the corresponding nadirs and peaks detected for calculation of MAGE:

### Glycemic Control Indicators

Glycemic Control Indicators were assessed to evaluate the effectiveness of diabetes management strategies by examining specific aspects of glucose control. The following metrics were included in the analysis:

- Fasting Glucose Proxy (FGxP) [16]: This metric estimates the fasting glucose levels based on the CGM data. It serves as a proxy for assessing baseline glycemic control and helps to approximate the glucose levels typically observed during fasting periods.
- Glucose Management Indicator (GMI) [29]: The GMI provides an estimate of average glucose levels by applying a formula that correlates with hemoglobin A1C values. This metric offers a standardized measure of overall glycemic control and helps in evaluating the long-term efficacy of diabetes management.
- Hypoglycemia events: This metric quantify the numbers of hypoglycemic episodes below 70 mg/dL for a minimum duration of 15 minutes.

Several methodologies have been proposed for approximating fasting glucose levels from CGM data. We employed a robust approach developed by Millard et al [16]., which does not require information about mealtimes. This method estimates fasting glucose by calculating the mean of the 30 lowest consecutive minutes of glucose readings during nighttime. *Figure 3* illustrates the CGM readings used to estimate fasting glucose from an individual.

**Figure 3.**
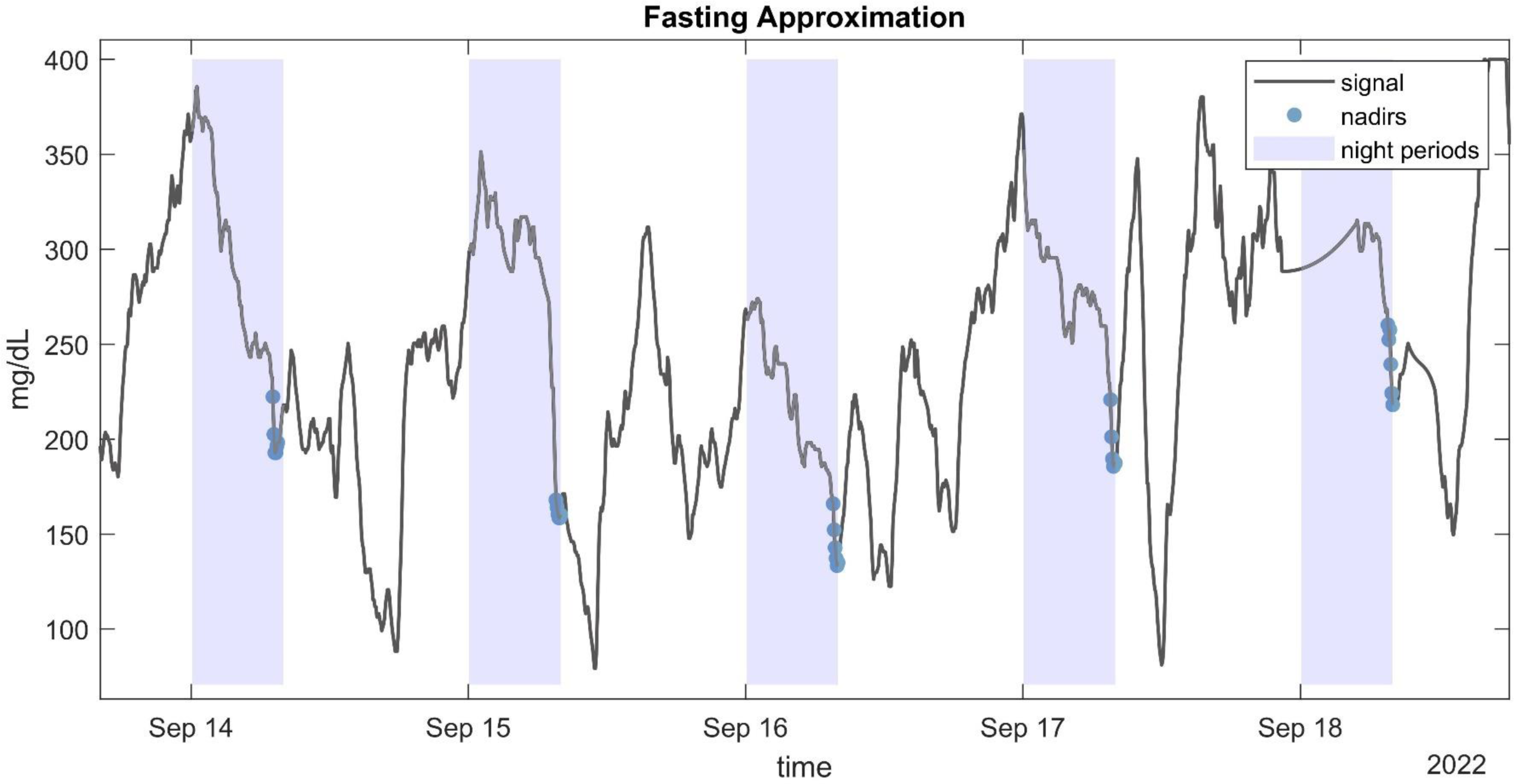
example of CGM signal with highlighted detected nadirs values during nights used for fasting approximation (FGxP).

GMI is calculated based on Bergenstal et al. [29]:

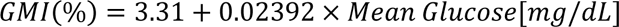

### Entropy and Complexity Measures

The entropy and complexity measure Multiscale Entropy (MSE) were utilized to evaluate the irregularity and unpredictability of glucose fluctuations, providing insights into the dynamic nature of glycemic control. MSE quantifies the complexity of the glucose time series by analyzing its entropy across multiple time scales. This metric captures the degree of randomness and structure within glucose data, providing a comprehensive measure of variability and the underlying complexity of glycemic fluctuations. The calculation involves embedding the data into various time scales and assessing the entropy at each scale. The sum of these entropy measures gives the Multiscale Entropy index, reflecting the overall complexity of the glucose signal. We adopted the implementation described by Kohnert et al. [30] where Multiscale Complexity Index (MCI) is defined as the sum over the range of scales, from 1 to 7, at the window length m = 2, the sensitivity criterion r = 0.15 times the standard deviation.

Tip: To reduce computational time significantly, the calculation of this metric can be omitted by removing the MSE code part.

## Example of usage

In this section, we demonstrate the application of QoCGM by deriving metrics from CGM data collected from individuals with Type 2 Diabetes Mellitus (T2DM) undergoing insulin therapy. We investigate the correlations between the various metrics generated by QoCGM.

### Study sample

We used CGM data from the *Diabetes teleMonitoring of patients in insulin Therapy (DiaMonT) trial (NCT04981808)*, comprising 331 people with insulin-treated T2DM [31]. We encompassed participants from both the intervention and the control arm. For consistency, we limited the analysis to the initial seven days of CGM monitoring post the inclusion date for all participants.

### Analyses

We conducted a coefficient of determination analysis (R^2^) to explore the linear relationships between the derived CGM metrics. R^2^ represents the proportion of variance in one metric that is explained by another metric. R^2^ ranges from 0 to 1, where 1 indicates that the variability in one variable can be completely explained by the other, and 0 indicates no explanatory power. This analysis provides insights into how different CGM-derived metrics are interrelated, offering guidance on their potential differences.

## Results

A total of 324 participants with T2DM had continuous glucose monitoring (CGM) data available for the first week following inclusion. The median (25th; 75th percentile) glucose level of the participants was 162 mg/dL (145; 193 mg/dL), with a time-in-range (TIR) of 65% (39; 78%), and a coefficient of variation (CV) of 24% (21; 29%). The coefficient of determination (R²) analysis is illustrated in a heatmap, *Figure 4*. Furthermore, coefficient of determination matrix plots with annotation of value is provided in *Supplementary Material S2*.

**Figure 4.**
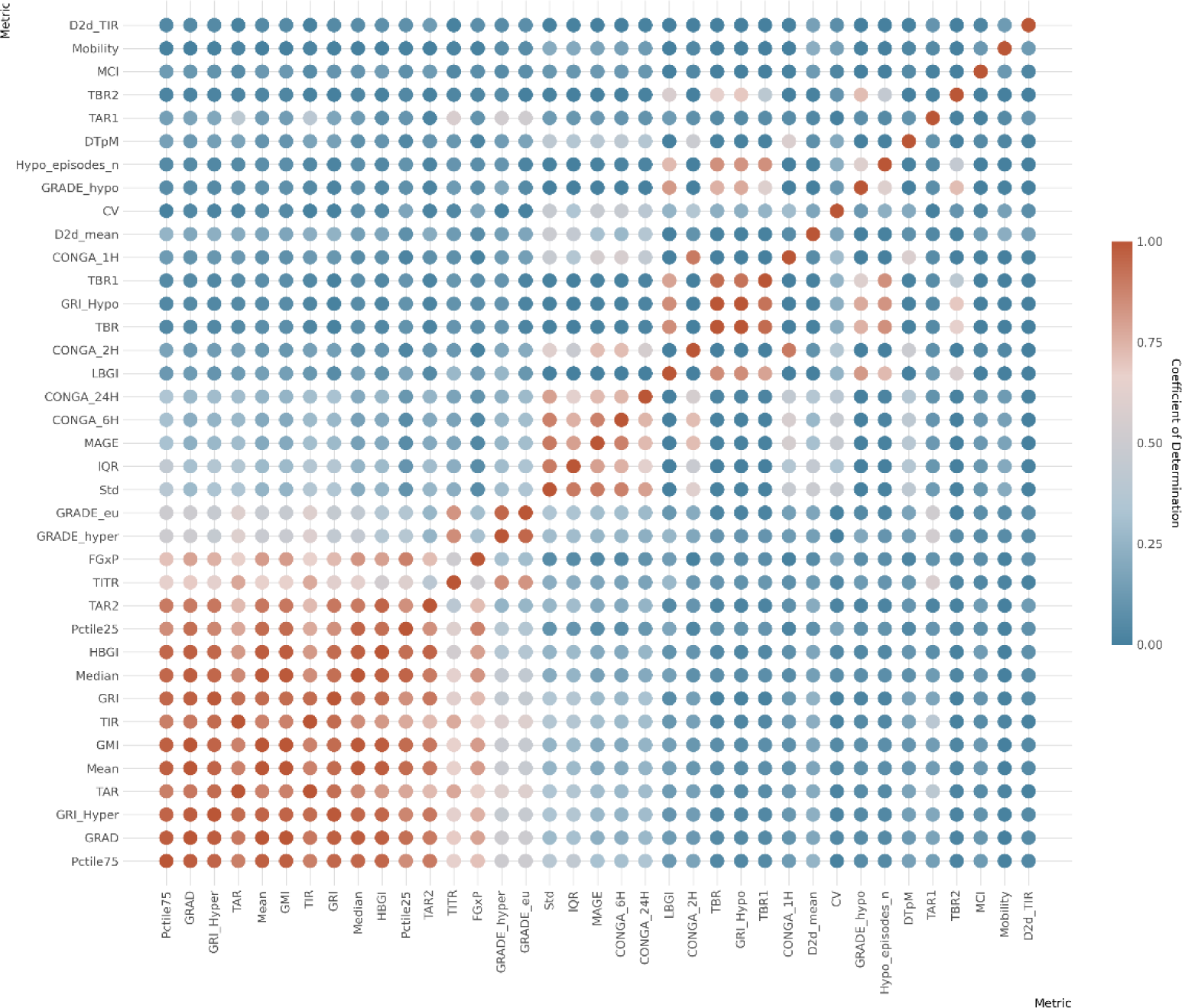
correlation heatmap for whole data CGM metrics. Metrics is arranged (top to bottom) with the lowest cumulative R² values across all metrics at top and the highest cumulative R² values at the bottom of the y-axe.

This analysis indicates that many metrics exhibit significant overlap with one or more other metrics. Notably, a few metrics from the whole-signal category show a low R² value (<0.5) when compared with other metrics. These include the day-to-day standard deviation of TIR and mean, mean continuous improvement (MCI), Mobility, and DTpM. *Figure 5* illustrates the six metrics with the lowest cumulative R² values across all metrics. For each of these metrics, the corresponding highest R² values are depicted in a circular plot. The analysis of diurnal and nocturnal metrics (*Supplementary Material S2)* also highlights significant differences between TIR metrics calculated from diurnal and nocturnal periods, underscoring the importance of measuring these as distinct entities in scientific studies.

**Figure 5.**
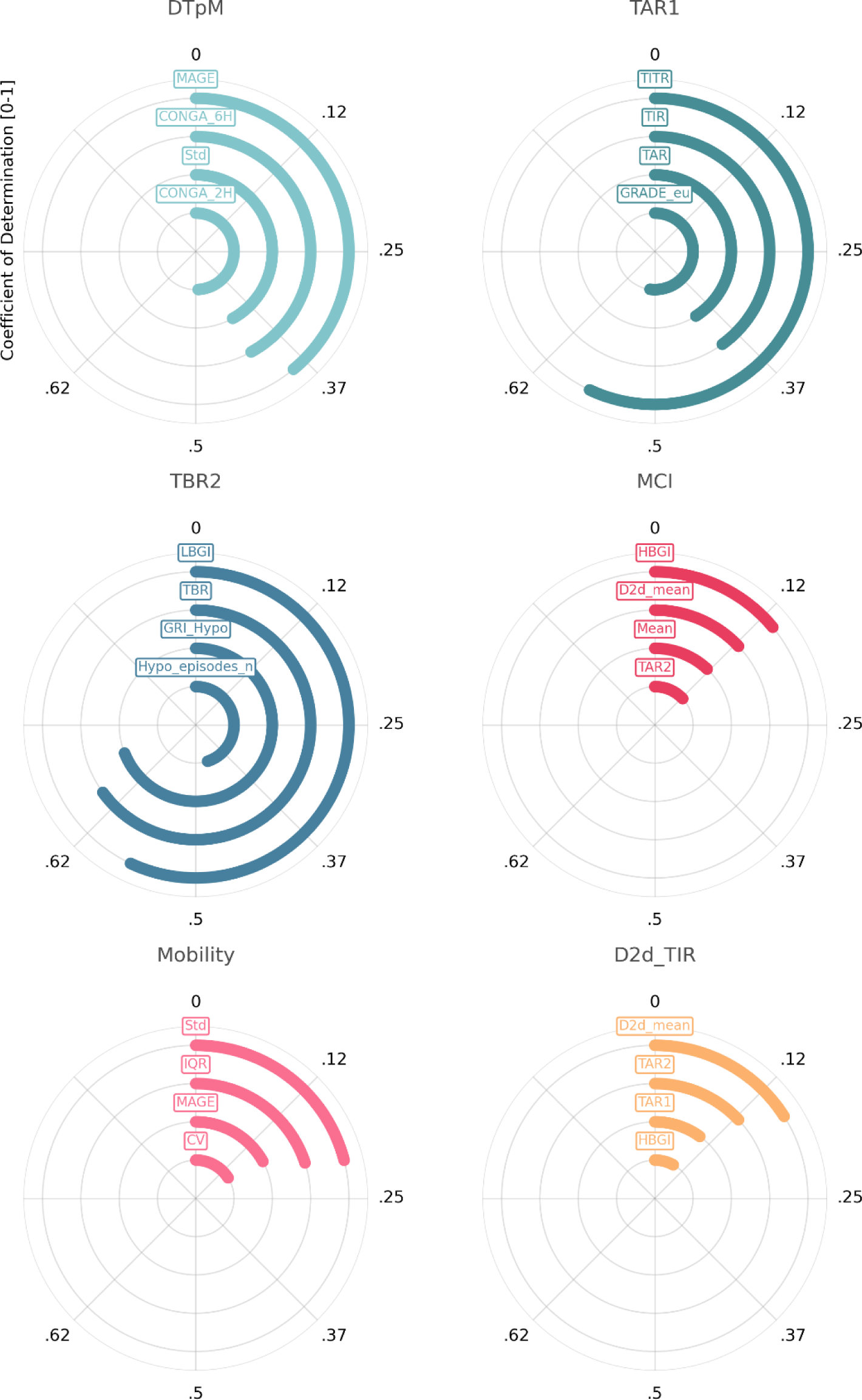
This figure illustrates the six metrics with the lowest cumulative R² values across all metrics. For each of these metrics, the corresponding highest R² values are depicted in a circular plot.

## Conclusions

In this paper, we introduced QoCGM, an open-source tool designed for researchers working with CGM data. QoCGM automates the preprocessing of data and derives a comprehensive set of metrics that capture key characteristics of glucose dynamics. The widespread adoption of this tool across various research populations could facilitate the identification of CGM metrics with the greatest clinical relevance in different contexts and patient groups. This can enhance the efficient and effective use of CGM in clinical research and practice. Additionally, a case study analyzing the coefficient of determination (R²) between the derived CGM metrics in people with insulin-treated T2DM demonstrated that different metrics capture distinct aspects of glucose dynamics. Notably, certain metrics exhibit a low R² with all other metrics, suggesting that they capture unique, independent information. Further research is required to explore the clinical implications of these findings.

## Abbreviations

CV: Coefficient of Variation
CSV: comma-separated value
CGM: Continuous Glucose Monitoring
CONGA: Continuous Overall Net Glycemic Action
DiaMonT: Diabetes teleMonitoring of patients in insulin Therapy
DTpM: Distance Traveled per Minute
FGxP: Fasting Glucose Proxy
GMI: Glucose Management Indicator
GRADE: Glycemic Risk Assessment in Diabetes
GRI: Glycemic Risk Index
IQR: Interquartile Range
LBGI/HBGI: Low/High Blood Glucose Index
MAGE: Mean Amplitude of Glycemic Excursions
PCHIP: Piecewise Cubic Hermite Interpolating Polynomial
QoCGM: Quantification of Continuous Glucose Monitoring
SD: Standard Deviation
TAR: Time-above-range
TBR: Time-below-range
TIR: Time-in-Range

## Acknowledgments

n/a.

## Author Contributions

SLC had access to all data analyzed in this study. SLC takes responsibility for the integrity and accuracy of the study data analysis and results (conceptualization, methodology, writing - Original Draft). Other authors were involved in data collection, writing - review & editing. All authors has approved the final manuscript.

## Funding received

the study on the QoCGM development was funded by i-SENS, Inc (Seoul, South Korea).

## Data Availability Statement

The data utilized in this study are not publicly available due to the inclusion of sensitive patient information, which is subject to strict confidentiality and privacy regulations. Access to the data is restricted to ensure compliance with ethical guidelines and to protect patient privacy. Requests for additional information or collaboration may be considered on a case-by-case basis, subject to appropriate ethical approval and data-sharing agreements.

