## Supplementary S1 for "From Data to Insights: A Tool for Comprehensive Quantification of Continuous Glucose Monitoring (QoCGM)"

### Supplementary material S1

Supplementary material S1 provide detailed explanation and mathematical definition of the CGM derived metrics.

MAGE — Identify significant excursions $\lambda$ above a threshold $\sigma$.

$$MAGE = \frac{1}{n}\sum\nolimits_{i=1}^{n} |\lambda_i| \; if \; (|\lambda_i| \geq \sigma)$$

GRI

$$GRI_{hypo} = \text{VLow} + (0.8 \cdot \text{Low})$$
$$GRI_{hyper} = \text{VHigh} + (0.5 \cdot \text{High})$$
$$VLow = \frac{1}{n}\sum\nolimits_{i=1}^{n} \begin{cases} 1 & if \; x_i < 54 \; mg/dL \\ 0 & otherwise \end{cases}$$
$$Low = \frac{1}{n}\sum\nolimits_{i=1}^{n} \begin{cases} 1 & if \; x_i < 70 \; \wedge x_i \geq 54 \; mg/dL \\ 0 & otherwise \end{cases}$$
$$\text{VHigh} = \frac{1}{n}\sum\nolimits_{i=1}^{n} \begin{cases} 1 & if \; x_i > 250 \; mg/dL \\ 0 & otherwise \end{cases}$$
$$High = \frac{1}{n}\sum\nolimits_{i=1}^{n} \begin{cases} 1 & if \; x_i \leq 250 \; \wedge x_i > 180 \; mg/dL \\ 0 & otherwise \end{cases}$$

Time in range

$$TIR = 100 \cdot \frac{1}{n}\sum\nolimits_{i=1}^{n} \begin{cases} 1 & if \; x_i \geq 70 \wedge x_i \leq 180 mg/dL \\ 0 & otherwise \end{cases}$$

Mobilty

$$Mobility = \sqrt{\frac{\text{variance}\left(\frac{\Delta x_i}{\Delta t}\right)}{\text{variance}\,(x)}}$$

$$\Delta x_i = x_{i+1} - x_i \; for \; i = 1,2, \ldots, n-1$$

CONGA

$$CONGA = \sqrt{\frac{\sum_{i=1}^{n}(x_i - \bar{x})^2}{n-1}}$$

LBGI
HBGI — Transform the glucose values:

$$f(BG) = 1.509 \times (\log(BG)^{1.084}) - 5.381$$

Calculate the risk function:

$$r(BG) = 10 \times \left(f(BG)\right)^2$$

Determine the LBGI:

$$LBGI = mean(r(BG) \times 1\{f(BG) < 0\})$$

Determine the HBGI:

$$HBGI = mean(r(BG) \times 1\{f(BG) > 0\})$$

GRADE

Calculate the GRADE score for each glucose value:

$$GRADE_i = 425 \times (log10(log10(\frac{BG_i}{18})) + 0.16)^2$$

Overall GRADE score:

$$GRADE = \frac{1}{n}\sum_{i=1}^{n} GRADE_i$$

GRADE for Hypoglycemia:

$$GRADEhypo = 100 \times \frac{\sum_{BG_{i<90}} GRADE_i}{GARDE}$$

DTpM

$$DTpM = \frac{\sum_{i=1}^{n-1} |BG_{i-1} - BG_i|}{n \times fs}$$
