## Supplementary S2 for "From Data to Insights: A Tool for Comprehensive Quantification of Continuous Glucose Monitoring (QoCGM)"

### Supplementary material S2

Supplementary material S2 provide annotations on $R^2$ between metrics. Annotated correlation matrix for (A) whole data metrics and (B) diurnal and nocturnal metrics. The annotations represent the R2 coefficient of determination, which indicate the proportion of variance in one metric that is explained by the other metrics.

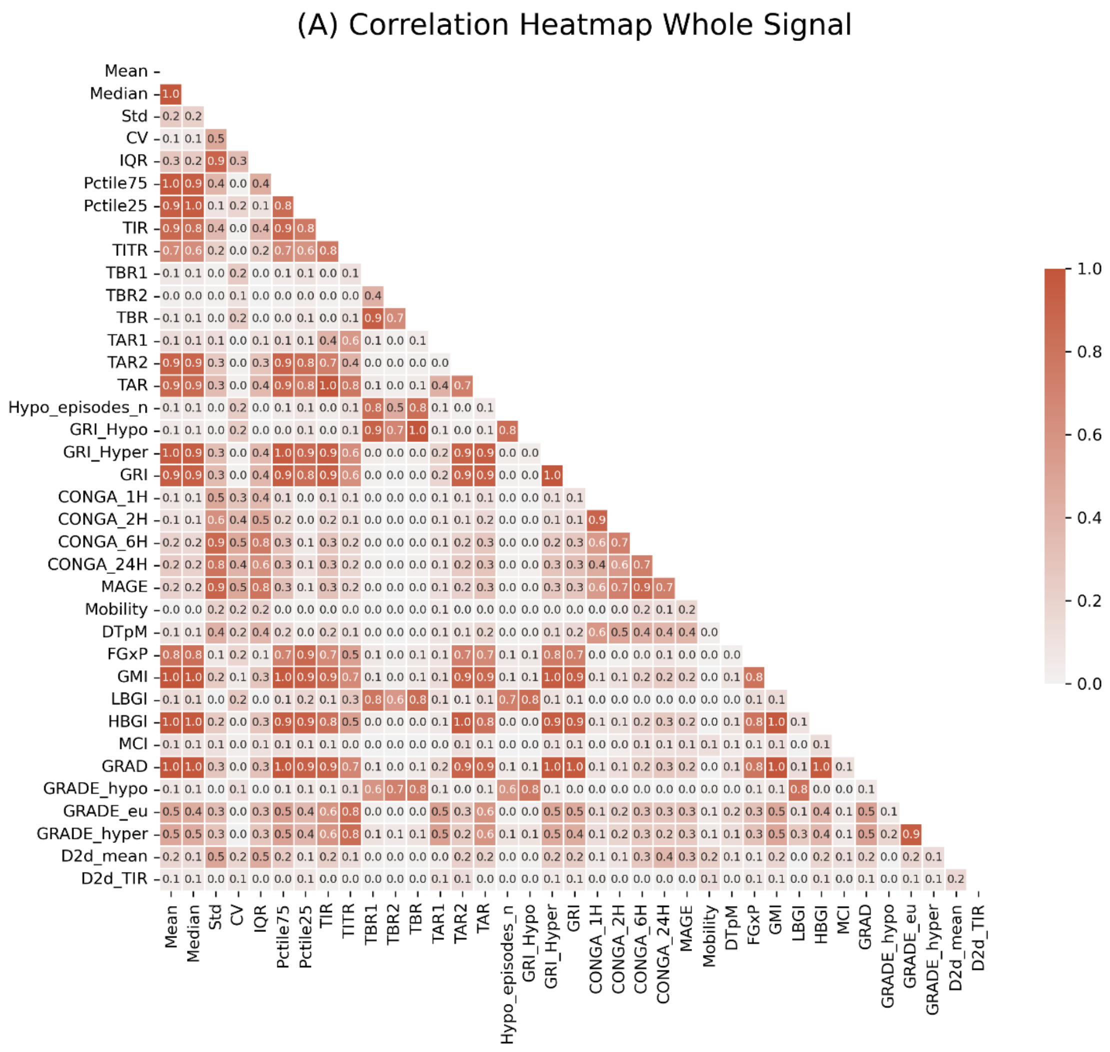

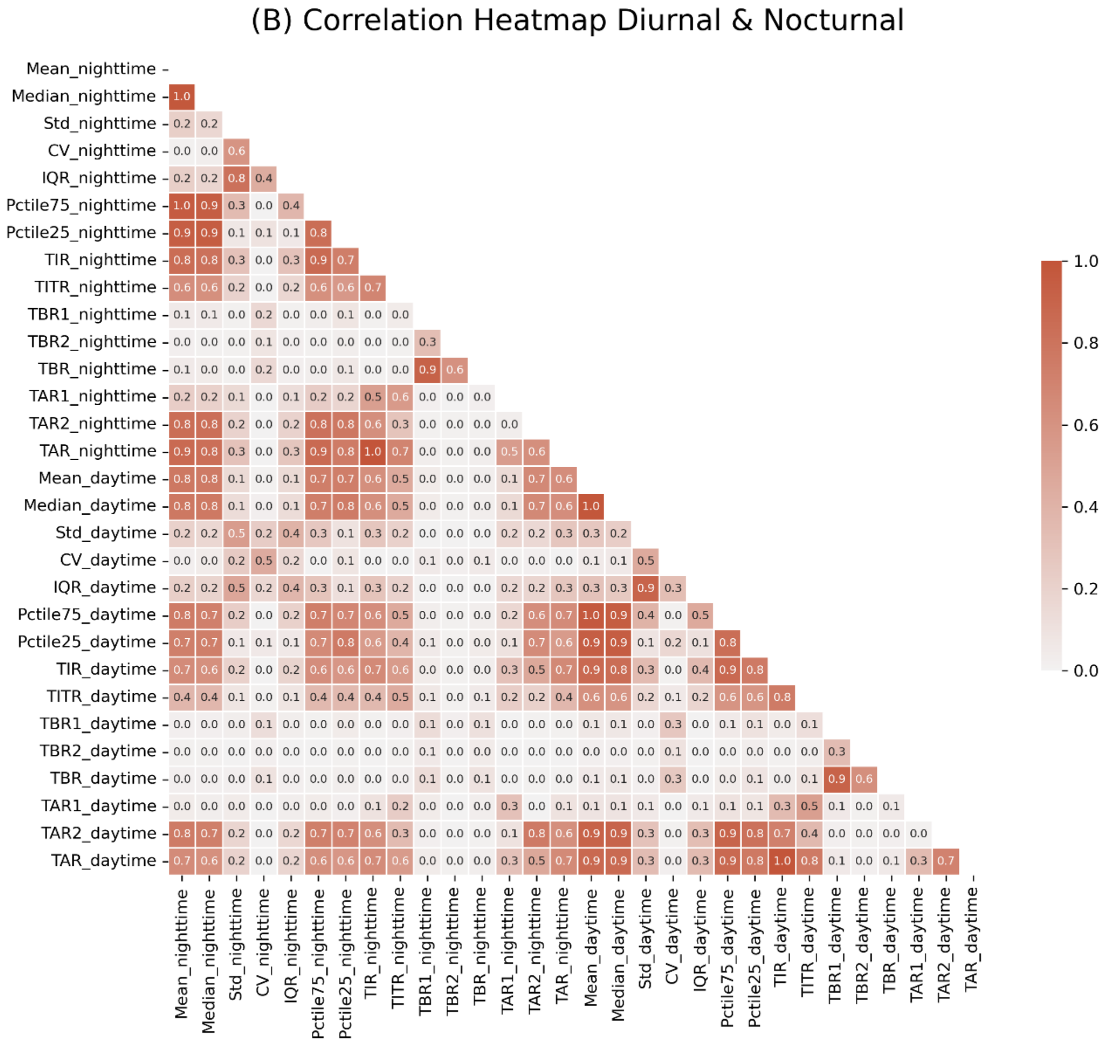

(B) Correlation Heatmap Diurnal & Nocturnal
